# Adaptation of the 20-Item Prosopagnosia Index for the screening of developmental prosopagnosia in Mexico

**DOI:** 10.1101/2025.09.10.25335477

**Authors:** Manuel Alejandro Mejía, Agustín Cardoso, Vania Lozoya, Antonieta Bobes-Leon

## Abstract

Developmental prosopagnosia (DP) is a lifelong condition characterized by difficulties in recognizing faces despite intact vision and intelligence. The 20-Item Prosopagnosia Index (PI20) is a widely used self-report instrument for screening face recognition deficits. However, no standardized Spanish version of the PI20 existed prior to this study, limiting its applicability in Spanish-speaking populations. This study aims to adapt and validate the PI20 for use in Mexican Spanish, ensuring its linguistic, cultural, and psychometric applicability for the Mexican population. Study 1 involved a rigorous translation and cultural adaptation process, including forward translation by multiple independent translators, expert panel review, and cognitive interviews with bilingual Mexican participants. Qualitative analysis of these interviews identified semantic and conceptual issues in several items, which were subsequently revised. A pilot test with 15 participants confirmed the clarity and feasibility of the adapted version, supporting its content validity. Study 2 evaluated the psychometric properties of the adapted PI20 in a sample of 333 adults. The results showed high internal consistency (McDonald’s omega and Cronbach’s alpha = .84) and good test–retest reliability (ICC_2,1_= .81). Confirmatory factor analysis supported a unidimensional structure consistent with the original scale, while exploratory analysis revealed two correlated components that did not undermine the overall model. The adapted PI20 showed a moderate negative correlation with the Cambridge Face Memory Test (r=-.229, p<.001), confirming convergent validity, and no significant association with the Cambridge Car Memory Test (r=-.106, p=.061), supporting discriminant validity. These findings demonstrate that the Mexican Spanish version of the PI20 is a reliable and valid tool for assessing DP in Spanish-speaking populations. This culturally adapted instrument can facilitate screening in clinical and research settings and contribute to a better understanding of face recognition difficulties across diverse linguistic and cultural groups.

## Introduction

Prosopagnosia, or “face blindness,” is a neurological disorder characterized by the difficulty or inability to recognize familiar faces. This condition can be present from birth, also known as developmental prosopagnosia (DP), or acquired as a consequence of brain injury (Garrido et al., 2008). The DP can have significant repercussions on the social and interpersonal lives of those affected (Corrow et al., 2016). The prevalence of DP has been estimated between 0.93% and up to 3.08% in samples of adults from the United States (DeGutis et al., 2023). However, these findings are not globally representative, and there are currently no prevalence data available for the Mexican population.

Furthermore, the study of DP has proven useful to understand face perception more generally. There are important theoretical questions about the basic mechanisms that allow face recognition, and whether those mechanisms are specialized for faces. The case study of DP has provided convergent evidence that face perception relies on the global integration of features rather than isolated analysis (Corrow et al., 2016). Although some individuals with DP experience additional difficulties with object or body recognition (Brewer et al., 2018; Stumps et al., 2020), evidence suggests that the core deficit predominates in facial identity processing, unlike with perception color , often remaining intact (Smith & Susilo, 2021). The current evidence suggest that DP is associated with cortical thinning in key areas of the face recognition network, in particular the fusiform face area (FFA) and the occipital face area (OFA), for the holistic processing of facial information (Anzellotti & Caramazza, 2014; Towler et al., 2017).

Proper assessment and diagnosis of this condition are essential to understanding its impact on individuals’ daily functioning and to developing interventions that address their specific needs. In this regard, various tools have been employed. Current instrument of assessment for diagnosis of DP suggest that a proper assessment should include tests of face perception such as the *Cambridge Face Perception Test* or the *Benton Facial Recognition Test* developed by Benton et al., (1994), tests of memory for unfamiliar faces, such as the *Cambridge Face Memory Test* designed by Duchaine y Nakayama (2006), and tests of memory for famous faces. In addition, self-report measures of lifelong difficulties with face recognition are also used, for example the PI20; (Shah et al., 2015), as well as measures that verify that the face perception deficit is not better explained by other cognitive or perceptual deficits, for which measures like the Cambridge Car Memory Test (CCMT) are used. For the self-report measure, the most widely used scale is PI20, which correlates with other objective face recognition measures and has proven effective in identifying symptoms associated with prosopagnosia (Shah et al., 2015).

The PI20 has been adapted into several languages—such as Japanese, French Danish , Chinese, Italian, and Portuguese (Nakashima et al., 2020; Nigrou et al., 2024a; Nørkær et al., 2023; Sun et al., 2021; Tagliente et al., 2023; Ventura et al., 2018a)—, but there is not a standardized Spanish version. This shortcoming limits its application in Spanish-speaking contexts, where cultural and linguistic norms may influence face perception and evaluation. In the Spanish-speaking world, and specifically in Mexico, there are few studies on prosopagnosia and limited adapted diagnostic tools, highlighting the need for validated instruments in these contexts.

The present study aims to validate the Mexican Spanish version of the PI20 with a mixed-methods design to provide a reliable and culturally appropriate instrument for the evaluation of developmental prosopagnosia, in accordance with international standards (Arizpe et al., 2019; Hambleton & Lee, 2013). This process involves translating the scale from British English into Spanish, comparing the translated version with the original, and evaluating its semantic and conceptual validity through expert feedback, cognitive interviews, and a pilot test. The objective was to ensure that the instrument is clear, culturally relevant, and reliable for assessing DPin the Spanish-speaking population. Finally, the procedure entailed evaluate the psychometric properties of the Spanish version of the PI20.

## Validation Study 1: Adapted Version of the PI20, and Evidence of Content Validity

### The first study

aimed to construct an initial Spanish version of the PI20 by following current guidelines of test adaptation, with the objective of producing a culturally appropriate version. The translation was carried out by independent translators, evidence to support content validity was produced from interviews with a variety of experts as well as cognitive interviews.

### Methods

#### Participants

Three different sets of participants were recruited. The first group was composed of three professional translators with Spanish as their native language and professional fluency in English. These translators worked independently from each other to ensure that the initial translation would reflect different perspectives and reduce potential bias. The second group of participants was selected to take part in cognitive interviews. This group consisted of five adult individuals, two men and three women, aged between 25 and 35 years, bilinguals (L1: Spanish, L2: English), and residents of Tijuana, Mexico. Recruitment for this group was carried out through digital diffusion strategies, such as online announcements and social media posts. Their role was to provide feedback on comprehension and interpretation of the items. Finally, the third group was recruited to complete a pilot version of the Spanish PI20. For this stage, 15 additional participants between 20 and 55 years of age were recruited at a local university to assess the clarity of instructions, the overall response time, and the internal consistency of the preliminary version of the instrument. The data obtained with the third group guided final adjustments before proceeding to the large-scale psychometric validation of Study 2.

#### Instruments

The primary instrument is the PI20 (Shah et al., 2015), a 20-item questionnaire designed to identify difficulties in facial recognition through a Likert scale ranging from 1 (“strongly disagree”) to 5 (“strongly agree”). The total score, which ranges between 20 and 100, is expected to reflect the degree of perceived difficulty, with scores above 65 being indicative of possible prosopagnosia. Previous studies have shown that the PI20 has high internal consistency (Cronbach’s alpha between 0.82 and 0.88; McDonald’s omega ≈ 0.81) and evidence of convergent validity—correlating with instruments such as the Cambridge Face Memory Test (CFMT)—as well as discriminant validity, demonstrated by the lack of correlation with tests assessing object recognition, such as the Cambridge Car Memory Test (CCMT) (Nakashima et al., 2020; Nigrou et al., 2024; Nørkær et al., 2023; Shah et al., 2015; Sun et al., 2021; Ventura et al., 2018).

#### Procedure

The adaptation process of the PI20 to Mexican Spanish followed the ITC guidelines ITC (International Test Commission, 2017) rigorously. A cross-sectional, descriptive, mixed-methods design was employed, integrating both quantitative data— obtained from the scores of the adapted PI20—and qualitative data, collected through cognitive interviews ( as per the recommendation by Sexton et al., 2023) that helped identify potential misunderstandings, ensuring conceptual equivalence.

Initially, three independent translators generated preliminary versions, which were then reconciled in a consensus meeting with a panel of four experts. The expert panel was composed of a specialist with experience in psychometrics and training in neuropsychology (this study’s first author), a researcher with extensive research in prosopagnosia and facial recognition (this study’s last author), and two experts with recognized experience in Mexican cultural contexts and trained in psychometrics (mentioned in the acknowledgements). The first PI20 Spanish version was presented back to the independent translators, who provided agreements and suggestions. Both the expert panel and the cognitive interviews offered qualitative feedback to refine items that had no initial consensus among translators (notably items 8, 18, and 20).

Cognitive interviews were conducted utilizing the initial Spanish version of the PI20, which was developed based on consensus from our group of expert translators to ensure the comprehension and cultural relevance of items within the instrument. These interviews helped identify items that could cause confusion in participants or misunderstandings deviating from the original meaning of said items, which subsequently supported the evaluation of the overall clarity of the questionnaire. Participants explained their interpretations of the items in a semi-structured format, where they explained whether they encountered any difficulties answering in detail.

Subsequently, a pilot study with 15 participants was carried out to evaluate response time, the clarity of instructions, as well as the internal consistency of the questionnaire, serving as the basis for final adjustments for Mexican cultural context.

#### Ethical Considerations

The study was conducted in accordance with the principles of the declaration of Helsinki (“World Medical Association Declaration of Helsinki,” 2013). All participants signed written informed consent, and the research protocol was approved by the authors institutional ethics committee (approval number #CEI-154), ensuring confidentiality and the proper use of the collected data.

### Results

The analyses revealed the need to adjust certain terms and phrases to improve comprehension in the Mexican context. These findings are summarized in Table 1.

**Table 1.** Findings from Cognitive Interviews on the Spanish Adaptation of the PI20.

| Items | Findings | Actions |
| --- | --- | --- |
| 1 to 7 | The first seven items were well understood by the participants, especially those related to self-assessment of the ability to recognize familiar faces and memorize faces, which presented little difficulty. The wording was considered clear and accessible. | Although they were generally well understood, some minor wording details were reviewed to optimize fluency and ensure there were no ambiguities. |
| 8 | Most participants adequately understood the intention of the item as the ability to generate a mental image of a person's face. However, a few isolated cases interpreted "imaginarme" as recalling past experiences, which evidenced the need to retain clear wording that would orient toward visual imagery. | The verb "imaginarme" was retained in the final wording to remain faithful to the original intention of the instrument, which seeks to assess the ability to mentally visualize faces and not the memory of past events or personal associations. |
| 9 to 17 | These items were mostly well received by the participants, with some minor observations about the wording. However, participants did not report significant comprehension issues. | The items were considered to be aligned with the participants' daily experiences, and no major adjustments were required. |
| 18 | Participants expressed general comprehension of this item, although with different interpretations of the term "confundir" (faces, names or identities). Responses showed that the formulation was functional in a family context and reflected real experiences, especially in large gatherings. | The wording "A veces confundo a los miembros de mi familia cuando nos reunimos" was retained, as it was adequately understood by the majority and retains the intent of the original construct. |
| 19 | Participants understood the item and successfully generalized it to real experiences, such as seeing before and after comparisons of celebrities prior to their fame. They mentioned concrete examples (e.g. celebrities that have undergone surgeries) and, some pointed out that certain changes make recognition difficult, in general they considered the statement clear and relevant. | The version: "Me resulta fácil reconocer a celebridades en imágenes de 'antes que fueran famosos' aunque hayan cambiado considerablemente" was retained, as it was correctly understood and adequately reflects the construct assessed. |
| 20 | Participants understood the item and related it to situations in which they see people they know outside their usual context. Although most of them indicated that they do not have difficulties in these cases, they recognized the scenario as a possible and clear experience. | The wording "Se me complica reconocer a personas conocidas cuando las veo fuera de situaciones o lugares habituales (ej., al encontrarme inesperadamente a un compañero de trabajo al hacer compras)" was maintained, as it was adequately understood and reflects the construct assessed. |

The cognitive interviews helped reach a consensus on items 8, 18, and 20, which had not been achieved during the initial translation phase between the independent translators. The results of the cognitive interviews resolved ambiguities and the translations were improved, ensuring their suitability for the Mexican context. These data can be seen in Table 2.

**Table 2.** Consensus on Items 8, 18, and 20 of the Spanish Adaptation of the PI20.

| Items | Findings | Actions |
| --- | --- | --- |
| **8: "I find it easy to picture individual faces in a crowd"**<br>Proposed translation: "Se me facilita imaginarme el rostro de alguien en medio de una multitud." | During translation, no consensus was reached on the use of "alguien" as opposed to terms such as "individuos". Some translators proposed more specific alternatives, although without clear agreement. | The item was reformulated as: "Se me facilita imaginarme el rostro de alguien en mi mente", as this version was more natural, general and understandable for the participants during the cognitive interviews. |
| **18: "At family gatherings, I sometimes confuse members of my family with others"**<br>Proposed translation: "A veces confundo a los miembros de mi familia con otras personas en reuniones familiares." | Although the sentence was considered clear by the translators, cognitive interview participants held different interpretations of the verb "confundir" (faces, names or identities), as a result, it was suggested to simplify the structure. | The final version was adjusted to: "A veces confundo a los miembros de mi familia cuando nos reunimos", as it was considered clearer and more in line with habitual Mexican Spanish usage, maintaining the intention of the original item. |
| **20: "It is hard to recognize familiar people when they are out of context"**<br>Proposed translation: "Se me complica reconocer a personas conocidas fuera de su contexto habitual." | Some translators expressed doubts about the clarity of the phrase "fuera de contexto" and suggested specifying the setting. Cognitive interviews confirmed that the difficulty arose in the interpretation of unexpected or unusual settings. | A contextual example was incorporated to improve clarity: "Se me complica reconocer a personas conocidas cuando las veo fuera de situaciones o lugares habituales (ej. al encontrarme inesperadamente a un compañero de trabajo al hacer compras)" which facilitated their understanding during cognitive interviews. |

### Conclusion of Study 1

A Spanish version of the PI20 was produced following international guidelines, including independent translators, expert consensus, cognitive interviews, and pilot testing. The strength of this process lies in the use of cognitive interviews as a systematic method of analysis, which allowed for the detailed exploration of how participants understood, interpreted, and responded to each item. This qualitative approach ensured that potential ambiguities or cultural mismatches were identified and corrected in real time. The results strongly supported the cultural and linguistic suitability of the Mexican Spanish version of the PI20, as participants provided concrete feedback that led to the resolution of problematic items—particularly items 8, 18, and 20—thereby improving the clarity, contextual relevance, and semantic equivalence of the scale. Taken together, these procedures provide solid evidence of content validity and a strong foundation for Study 2 to estimate the psychometric properties of the instrument.

## Validation Study 2: Reliability and Evidence of Validity

The purpose of the second study was to gather evidence of reliability and construct validity of the Spanish version of the PI20. For this objective, a cross-sectional correlational study was conducted with a larger sample and the inclusion of additional face and object recognition tests. The study tested for evidence of internal structure, convergent validity with face perception and face memory, and discriminant validity with object memory.

### Methods

#### Participants

A total of 333 participants completed the Spanish version of PI20, including the 15 participants from the pilot in Study 1. The additional 318 participants recruited for Study 2 also completed a face recognition battery described below. Two participants did not complete the battery, but their data was included for tasks completed fully (one task for one participant, and two tasks for the other). The sample included 127 women (38.14%) and 206 men (61.86%), between 18 to 65 years old (M = 28.58, SD = 11.8). Participants were recruited through non-probabilistic convenience sampling, with efforts made to ensure diversity in terms of age, educational background and socioeconomic status (see Table 3).

**Table 3.** Sociodemographic characteristics of participants who completed the PI20 and the face recognition battery (Study 2, n = 333).

| Variables | M / F | SD / % |
| --- | --- | --- |
| Age in years | 28.58 | 11.8 |
| Gender |  |  |
| Women | 127 | 38.14 |
| Men | 206 | 61.86 |
| Education |  |  |
| No education | 2 | 0.6 |
| Primary | 4 | 1.2 |
| Secondary | 41 | 12.31 |
| High School | 206 | 61.86 |
| Undergraduate | 75 | 22.52 |
| Graduate studies | 5 | 1.5 |

*Note.* M = Mean, F = Frequency, SD = Standard Deviation

The sample size was estimated from power analyses. For the correlations between tests, power analyses run with the *pwr* package in R (Champely, 2020), in RStudio (Posit team, 2023), indicated that a sample size of at least 164 would have 90% power to detect effects of r > .25 with an alpha level of 0.05. For the evidence of internal structure, power analysis for a confirmatory factor analysis (CFA) indicated that a sample size of 147 would have 90% power to detect a root mean square error of approximation (RMSEA) of 0.05 for a model of a single factor with 20 items (using calculator by Arifin, 2025; that uses calculations from Kim, 2005).

For internal consistency and factor structure analyses, data from all 333 participants with complete responses across all PI20 items was used. Additionally, a subsample of 31 participants completed a second session after approximately six weeks to assess test–retest reliability.

#### Instruments and Procedure

The Mexican Spanish version of PI20 obtained from Study 1 was administered on a large scale under controlled conditions, alongside a standardized battery of face processing tests. This battery included the Cambridge Face Memory Test (CFMT), the Cambridge Face Perception Test (CFPT), and the Cambridge Car Memory Test (CCMT). These tasks were also adapted into Spanish by the authors of this study by translating the instructions and retaining the overall structure and stimuli. The tasks were implemented in PsychoPy (Peirce et al., 2019). CFMT and CFPT were used to assess convergent validity, while the CCMT was used to obtain discriminant validity of the PI20. This methodological approach allowed for a direct comparison between the self-report of the PI20 to face recognition performance and object memory abilities, strengthening the psychometric evaluation of the Spanish version of the PI20. Participants completed the PI20 and the standardized battery within Pavlovia (https://pavlovia.org/).

The study was conducted in full compliance with the ethical procedures established in Study 1. Data analyses were performed using JASP 0.19.3 Team, 2025.

### Results

#### Descriptive results of the face battery

Table 4 presents the descriptive statistics for PI20 and the face processing battery. The average total score on the PI20 was 43.68 (SD = 9.36), with higher scores indicating higher self-reported difficulty in face recognition. In the Cambridge Face Memory Test (CFMT), the mean score was 54.41 correct trials (SD = 10.26; N = 318; maximum score = 72). The Cambridge Car Memory Test (CCMT) had an average of 45.69 correct trials (SD = 9.44; N = 315; maximum score = 72), while the Cambridge Face Perception Test (CFPT), which is scored by total errors, showed a mean of 48.99 (SD = 20.99; N = 316).

**Table 4.** Descriptive statistics of the Spanish version of the PI20 and the Cambridge battery (CFMT, face memory; CCMT, car memory; CFPT, face perception) in Study 2.

| **Test** | **Mean** | **SD** | **Min** | **Max** | **N** |
| --- | --- | --- | --- | --- | --- |
| CFMT Total | 54.41 | 10.26 | 29 | 72 | 318 |
| CCMT Total | 45.69 | 9.44 | 23 | 72 | 315 |
| CFPT Total errors | 48.99 | 20.99 | 10 | 114 | 316 |
| PI20 Total | 43.68 | 9.36 | 21 | 70 | 333 |
| **PI20 Item totals** |  |  |  |  |  |
| PI20 Item 01 | 2.23 | 0.93 | 1 | 5 | 333 |
| PI20 Item 02 | 2.29 | 0.99 | 1 | 5 | 333 |
| PI20 Item 03 | 3.45 | 1.00 | 1 | 5 | 333 |
| PI20 Item 04 | 2.05 | 0.97 | 1 | 5 | 333 |
| PI20 Item 05 | 1.59 | 0.71 | 1 | 4 | 333 |
| PI20 Item 06 | 1.91 | 0.85 | 1 | 5 | 333 |
| PI20 Item 07 | 1.81 | 0.93 | 1 | 5 | 333 |
| PI20 Item 08 | 2.69 | 1.17 | 1 | 5 | 333 |
| PI20 Item 09 | 3.11 | 0.99 | 1 | 5 | 333 |
| PI20 Item 10 | 2.14 | 0.87 | 1 | 5 | 333 |
| PI20 Item 11 | 1.79 | 0.91 | 1 | 5 | 333 |
| PI20 Item 12 | 2.13 | 0.97 | 1 | 5 | 333 |
| PI20 Item 13 | 1.96 | 1.02 | 1 | 5 | 333 |
| PI20 Item 14 | 1.88 | 0.92 | 1 | 5 | 333 |
| PI20 Item 15 | 1.69 | 0.87 | 1 | 5 | 333 |
| PI20 Item 16 | 1.86 | 0.95 | 1 | 5 | 333 |
| PI20 Item 17 | 2.80 | 1.14 | 1 | 5 | 333 |
| PI20 Item 18 | 1.39 | 0.63 | 1 | 5 | 333 |
| PI20 Item 19 | 2.85 | 1.04 | 1 | 5 | 333 |
| PI20 Item 20 | 2.11 | 0.97 | 1 | 5 | 333 |

#### Reliability of the PI20

The Spanish version of PI20 demonstrated strong internal consistency and temporal reliability. The McDonald’s Omega and Cronbach’s Alpha coefficients for the full PI20 scale were both 0.84. Using the method proposed by Bonett and Wright (2015), the 95% confidence intervals of the Spanish version were estimated to range from 0.82 to 0.87 for Omega and from 0.81 to 0.86 for Alpha, indicating robustness of the results. The intraclass correlation coefficient (ICC2,1), used to assess test–retest stability, was 0.81(95% CI [0.762,0.844], n=31), further supporting the scale’s temporal robustness.

Comparative estimates of internal consistency for the face processing battery also indicated high reliability. The CFMT showed the highest internal consistency (ω = 0.91; α = 0.90), while the CCMT and CFPT demonstrated solid reliability as well (ω = 0.84 and 0.83; α = 0.84 and 0.82, respectively). The test-retest values for the face processing battery presented in Table 5 exceed the .5 threshold for all tests, values ranging from 0.66 (CFPT) to 0.75 (CCMT), indicating moderate to good intraclass correlation (Koo & Li, 2016).

**Table 5.** Reliability indices for the Spanish version of the PI20 (CFMT, face memory; CCMT, car memory; CFPT, face perception; Study 2).

| Test | Coefficient | Estimate | Lower | Upper | Std. Error |
| --- | --- | --- | --- | --- | --- |
| PI20 | McDonald's $\omega$ | 0.84 | 0.82 | 0.87 | 0.01 |
| | Cronbach's $\alpha$ | 0.84 | 0.81 | 0.86 | 0.01 |
|  | Mean | 43.68 | 42.67 | 44.68 | 0.51 |
|  | SD | 9.36 | 8.70 | 10.13 | 0.35 |
|  | ICC2,1 | 0.81 | 0.76 | 0.84 |  |
| CFMT | McDonald's $\omega$ | 0.91 | 0.89 | 0.92 | 0.01 |
| | Cronbach's $\alpha$ | 0.90 | 0.89 | 0.92 | 0.01 |
|  | Mean | 51.96 | 50.33 | 53.58 | 0.83 |
|  | SD | 0.92 | 0.92 | 55.50 | 1.00 |
|  | ICC2,1 | 0.68 | 0.18 | 0.85 |  |
| CCMT | McDonald's $\omega$ | 0.84 | 0.82 | 0.87 | 0.01 |
| | Cronbach's $\alpha$ | 0.84 | 0.82 | 0.87 | 0.01 |
|  | Mean | 45.69 | 44.65 | 46.73 | 0.53 |
|  | SD | 9.44 | 8.75 | 10.24 | 0.33 |
|  | ICC2,1 | 0.66 | 0.60 | 0.72 |  |
| CFPT | McDonald's $\omega$ | 0.83 | 0.80 | 0.85 | 0.01 |
| | Cronbach's $\alpha$ | 0.82 | 0.80 | 0.85 | 0.01 |
|  | Mean | 128.46 | 125.03 | 131.88 | 1.75 |
|  | SD | 31.04 | 28.79 | 33.66 | 1.13 |
|  | ICC2,1 | 0.75 | 0.70 | 0.80 |  |

Together, these results support the internal structural validity of the PI20 and confirm that its adaptation retains psychometric integrity when compared to established face and object recognition instruments, as shown in Table 5. Furthermore, as argued by Stensen & Lydersen (2022), McDonald’s omega may offer a more appropriate measure of internal consistency than Cronbach’s alpha, particularly when the assumption of tau-equivalence is questionable.

tem-level analyses of PI20 supported the internal reliability of the adapted scale. McDonald’s Omega coefficients ranged from 0.826 to 0.858, and Cronbach’s Alpha values ranged from 0.815 to 0.851 across the 20 items, indicating consistent item performance. Although the removal of items 8 and 19 produced marginal increases in reliability estimates, the improvements were not substantial enough to justify their exclusion. These results suggest that all items contribute significantly to the measurement of difficulties associated with developmental prosopagnosia. A detailed summary of the item-level reliability statistics is presented in Table 6.

**Tabla 6.** Reliability of the Spanish version of the PI20 if each item was deleted (Study 2).

| Item | McDonald's $\omega$ | Cronbach's $\alpha$ | Mean | SD |
| --- | --- | --- | --- | --- |
| PI20_01 | 0.829 | 0.824 | 2.225 | 0.932 |
| PI20_02 | 0.827 | 0.821 | 2.291 | 0.992 |
| PI20_03 | 0.848 | 0.846 | 3.447 | 0.997 |
| PI20_04 | 0.828 | 0.822 | 2.051 | 0.973 |
| PI20_05 | 0.829 | 0.824 | 1.586 | 0.709 |
| PI20_06 | 0.829 | 0.824 | 1.907 | 0.850 |
| PI20_07 | 0.821 | 0.816 | 1.805 | 0.932 |
| PI20_08 | 0.848 | 0.841 | 2.685 | 1.169 |
| PI20_09 | 0.841 | 0.835 | 3.105 | 0.990 |
| PI20_10 | 0.831 | 0.826 | 2.141 | 0.872 |
| PI20_11 | 0.829 | 0.823 | 1.790 | 0.911 |
| PI20_12 | 0.822 | 0.817 | 2.126 | 0.974 |
| PI20_13 | 0.839 | 0.831 | 1.955 | 1.018 |
| PI20_14 | 0.835 | 0.829 | 1.877 | 0.918 |
| PI20_15 | 0.829 | 0.823 | 1.691 | 0.870 |
| PI20_16 | 0.828 | 0.823 | 1.856 | 0.946 |
| PI20_17 | 0.846 | 0.840 | 2.796 | 1.143 |
| PI20_18 | 0.833 | 0.828 | 1.387 | 0.628 |
| PI20_19 | 0.850 | 0.845 | 2.847 | 1.043 |
| PI20_20 | 0.832 | 0.827 | 2.111 | 0.971 |

#### Factor analysis

Confirmatory factor analysis (CFA) was conducted to test the unidimensional structure of the Mexican Spanish version of PI20, as proposed in the original scale. The model, estimated by using diagonally weighted least squares as recommended for ordinal items (Li, 2016), had acceptable fit indices: the Comparative Fit Index (CFI) was .960, and the Tucker–Lewis Index (TLI) was .955, both exceeding the conventional cutoff of .90. The Root Mean Square Error of Approximation (RMSEA) was .089 (90% CI [.081, .096]), and the Standardized Root Mean Square Residual (SRMR) was .078, both near acceptable thresholds (Hu & Bentler, 1999)..

The results support the adequacy of a one-factor solution, confirming that the items of the PI20 measure a common underlying construct associated with developmental prosopagnosia. Most factor loadings were statistically significant and above the .30 threshold, while a few items (3, 8, 17, and 19) showed lower loadings (< .30). Nonetheless, all items contributed significantly to the model, supporting their retention in the scale. A summary of the model fit indices and parameter estimates is provided in Supplementary Table S2.

An exploratory factor analysis (EFA) was conducted to further examine the underlying structure of the Mexican Spanish version of PI20. The analysis revealed a two-factor solution using a principal axis factoring with oblique rotation. Items such as PI20_07 (*“En ocasiones tengo que advertirles a las personas nuevas que conozco que soy malo reconociendo los rostros*”) and PI20_12 (“*Tengo que esforzarme más que otras personas para memorizar rostros*”) loaded strongly on factor 1, whereas items such as PI20_08 (“*Se me facilita imaginarme el rostro de alguien en mi mente*”) and PI20_13 (“*Confío mucho en mi habilidad para reconocerme en fotografías*”) showed higher loadings on factor 2, suggesting the presence of two distinct but strongly related components within the DP construct. Items in factor 2 characterized as items where agreement with the sentence indicated better face recognition abilities. Factors were correlated (r=.330).

These results support the adequacy of the one-factor solution, confirming that the PI20 items measure a common underlying construct associated with developmental prosopagnosia. All factor loadings were statistically significant and above 0.28, indicating satisfactory contributions of individual items to the latent trait. A summary of the model fit indices and parameter estimates is presented in Table 7.

**Table 7.**
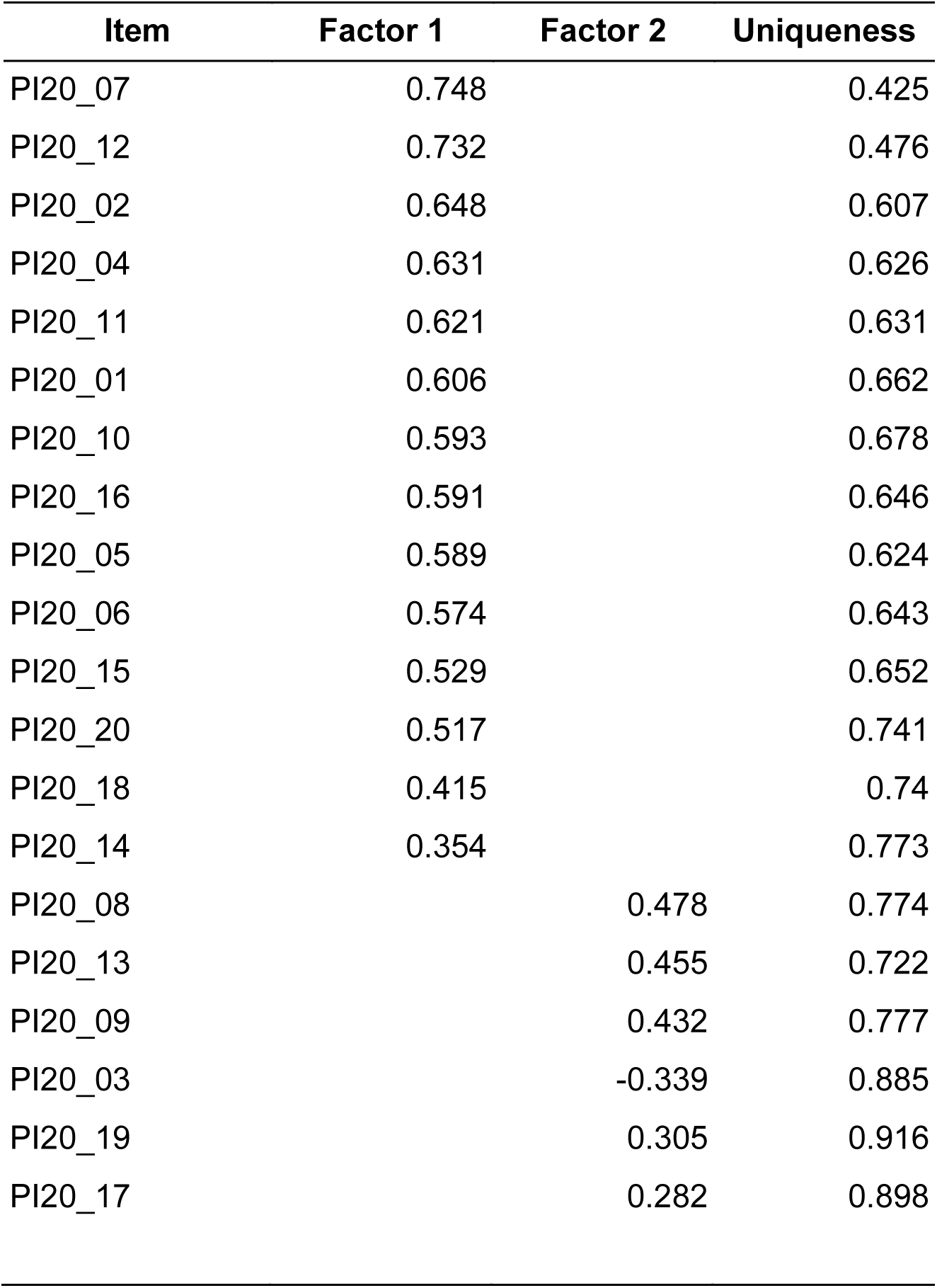
Exploratory Factor Analysis of the Spanish version of the PI20 (Study 2).

#### Convergent & discriminant Validity

Evidence for convergent and discriminant validity of PI20 was assessed through correlations with objective performance on face and object recognition tasks. As expected, the PI20 showed a significant negative correlation with the Cambridge Face Memory Test (CFMT; r = –.229, p < .001), indicating that individuals who reported greater difficulties in face recognition tended to perform worse on a standardized test of face memory. This correlation aligns with the threshold proposed by Gignac & Szodorai (2016), who consider effect sizes of r ≈ .22 to reflect moderate associations, thereby supporting the convergent validity of the instrument.

In contrast, the PI20 did not show a significant correlation with errors in the Cambridge Face Perception Test (CFPT; r = .047, p = .41), nor with its accuracy-based scoring (r = –.139, p = .08), providing limited support for convergent validity with perceptual tasks. No significant association was observed with the Cambridge Car Memory Test (CCMT; r = –.106, p = .061), supporting the discriminant validity of the PI20 in relation to non-face object memory. The CCMT was specifically designed to mirror the CFMT in format while assessing memory for non-face objects (cars), thus allowing for a controlled evaluation of discriminant validity (Dennett et al., 2012). These results confirm that the PI20 specifically reflects face recognition difficulties rather than general visual memory performance. All correlations are reported in Table 8.

**Tabla 8.** Correlations between the Spanish version of the PI20 with CFMT, CFPT and CCMT (CFMT, face memory; CCMT, car memory; CFPT, face perception; Study 2).

|  | **PI20_Total** | **CFMT_Total** | **CFPT_total_errors** | **CCMT_Total** |
| --- | --- | --- | --- | --- |
| PI20_Total | — |  |  |  |
| p-value | — |  |  |  |
| CFMT_Total | -0.229 | — |  |  |
| p-value | < .001 | — |  |  |
| CFPT_total_errors | 0.047 | -0.481 | — |  |
| p-value | 0.407 | < .001 | — |  |
| CCMT_Total | -0.106 | 0.283 | -0.257 | — |
| p-value | 0.061 | < .001 | < .001 | — |

### Conclusion of Study 2

The psychometric evaluation of the adapted Prosopagnosia Index 20 (PI20) instrument confirmed it is a valid and reliable instrument for the assessment of DPin the Mexican context. The instrument demonstrated high internal consistency and excellent test-retest reliability. The internal structure was consistent with the original unidimensional model. Convergent validity was supported by a negative correlation with the Cambridge Face Memory Test (CFMT); on the other hand, discriminant validity was established by having no correlations with the Cambridge Car Memory Test (CCMT). Items with low factor loadings do not significantly affect internal consistency or reliability, so there is no drawback to their inclusion. The results support, in general, PI20 as a useful instrument to identify difficulties in face recognition in Spanish-speaking contexts.

## General Discussion

This research successfully adapted and validated PI20 for use in Mexican Spanish for the screening of DP, following a rigorous, multi-phase process that ensured semantic, conceptual, and cultural equivalence. In Study 1, cognitive interviews and pilot testing with native Spanish speakers from Mexico enabled the refinement of specific items, particularly items 8, 18, and 20, to enhance clarity and contextual relevance. The results supported the content validity of the adaptation and laid a solid foundation for subsequent psychometric evaluation. In Study 2, the adapted instrument demonstrated robust psychometric properties in a larger sample. The PI20 showed high internal consistency, excellent test–retest reliability, and a factorial structure consistent with the original unidimensional model. Evidence of convergent validity was established through a negative correlation with the Cambridge Face Memory Test (CFMT), while discriminant validity was confirmed by the absence of significant correlations with the Cambridge Car Memory Test (CCMT), an object recognition task. Exploratory factor analysis suggested the presence of two correlated components, though the overall structure remained consistent with a unidimensional interpretation, as confirmed by confirmatory factor analysis.

These findings confirm there is adequate preliminary evidence of reliability and validity of the Mexican Spanish version of the PI20 for identifying difficulties associated with DPamong Spanish-speaking populations in Mexico. Its psychometric integrity, combined with its cultural and linguistic appropriateness, makes it a valuable tool for both clinical screening and research.

The adaptation process followed internationally recognized guidelines, including those from the (International Test Commission, 2017), and involved forward translation, expert committee review, and iterative refinement based on cognitive interviews and pilot testing. As in other adaptations, such as the Danish and French versions, cultural and linguistic challenges emerged—particularly in items requiring contextual reinterpretation (e.g., item 8)—and were resolved through methodologically rigorous procedures (Sexton et al., 2023). The development of culturally adapted diagnostic instruments is essential for advancing the assessment of neurocognitive conditions in diverse populations.

The use of a mixed-methods, cross-sectional design enabled a comprehensive evaluation of the instrument. In Study 1, qualitative insights from cognitive interviews provided content validity evidence, ensuring the clarity and cultural relevance of the translated items. In study 2, psychometric analyses based on large-scale administration confirmed the instrument’s structural, convergent, and discriminant validity. This methodological integration aligns with best practices in test adaptation and validation (Arizpe et al., 2019; Hambleton & Lee, 2013), resulting in a version of the PI20 that is both psychometrically sound and contextually appropriate.

The average scores of the Spanish version of the PI20 were highly similar to those reported by Shah et al. (Shah et al., 2015). Particularly, the mean PI20 score obtained in this study was comparable to their control sample (42 vs 38.90), and the pattern of the means for each item was also equivalent. For example, the items 3, 9, and 19 (3.69, 2.76, and 2.79, respectively), had the highest means in Shah et al. (2015), and also were the highest in our study (3.45, 3.11, and 2.85, respectively). Therefore, the descriptive results obtained with the Spanish version of the PI20 in the Mexican sample are also evidence of the equivalence to the original English version.

Reliability analyses revealed excellent internal consistency, with McDonald’s Omega and Cronbach’s Alpha both at .84—comparable to those of the original scale <u>(Shah et al., 2015)</u> and other adaptations (Nørkær et al., 2023). Item-level analysis showed no substantial increase in reliability upon the removal of any item, supporting the internal coherence of the scale. Test–retest reliability, assessed over a six-week interval, produced an intraclass correlation coefficient (ICC) of .81, consistent with standards for temporal stability (Koo & Li, 2016).

Construct validity was supported by both confirmatory and exploratory factor analyses, which indicated a predominantly unidimensional structure. This aligns with the theoretical basis of the PI20 and previous validation studies. Evidence for convergent validity was provided by a moderate negative correlation between PI20 scores and CFMT performance (r = –.229), matching the threshold for moderate effects proposed by Gignac and Szodorai (2016) and consistent with prior findings (Gray et al., 2017). Discriminant validity was demonstrated by the absence of a significant correlation with the Cambridge Car Memory Test (CCMT), confirming that the PI20 specifically measures face recognition difficulties rather than general object memory.

The weak and nonsignificant correlations observed between PI20 and the Cambridge Face Perception Test (CFPT) suggest that face perception and face memory may represent partially dissociable cognitive processes. This pattern is consistent with previous findings reported by DeGutis et al. (2023) and Nørkær et al. (2023), and supports the relevance of considering subtypes of developmental prosopagnosia—such as apperceptive and associative forms—in both research and clinical assessments.

The current adaptation of the PI20 contributes to this goal and paves the way for future studies exploring its applicability across broader demographic groups, including children, older adults, and clinical populations. Additionally, examining sociocultural variables that may influence face perception can further inform the contextualization of prosopagnosia and guide tailored intervention strategies. For example, prior studies, such as Nigrou et al., (2024, have found associations between higher PI20 scores and increased symptoms of social anxiety, implying that face recognition deficits may have broader psychosocial implications. Although such variables were not examined in the present study, they represent potential directions for future research within the Mexican population.

In conclusion, the Mexican Spanish version of The 20-Item Prosopagnosia Index (PI20) possesses promising initial evidence for its use as a culturally appropriate, reliable and valid tool for identifying developmental prosopagnosia. Its strong psychometric performance and sensitivity to cultural context make it a valuable instrument for both clinical applications and research in Spanish-speaking populations. Future studies may extend this work by exploring its utility across diverse age groups and by examining the psychosocial and cognitive correlates of face recognition deficits in greater depth.

## Data Availability

All the translation materials, anonymized participant data, and analysis scripts can be found on this paper's project page on the https://osf.io/k8vwt/?view_only=289b9c1da7684c3baffe29a8604d15a5

## Author contributions

Contributed to conception and design: MAM, AC, VL, ABL

Contributed to acquisition of data: AC, MAM, VL

Contributed to analysis and interpretation of data: MAM, AC, VL, ABL

Contributed to supervision, and data curation: MAM

Drafted and/or revised the article: MAM, AC, VL

Approved the submitted version for publication: MAM, AC, VL

## Acknowledgements

**Data collection**: Alam Ibarra, Javier Samaniego Ojeda, Emily Sayuri Morales Castelan, Azul Gabriela Camacho Hernandez, Perfecto Sanchez Maria de la Luz, & Velasco Lopez Ximena Estephanya. **Drafts revisions**: Alam Ibarra. **Translators**: Ricardo Eliud Ramirez, Genesis Cerrillo, & Esmeralda Mosqueda. **Expert panel**: Víctor Fuentes-Barradas & Diana Estefanía Andrade.

## Funding information

MM received funding from the Institutional Research Coordination (Project #517) of CETYS University.

## Ethics statement

Ethical approval was obtained from the Research Ethics Committee of CETYS University (#D-CEI154).

## Competing interests

Authors declare no competing interests for this study.

## Supplemental material

**Table S1.**
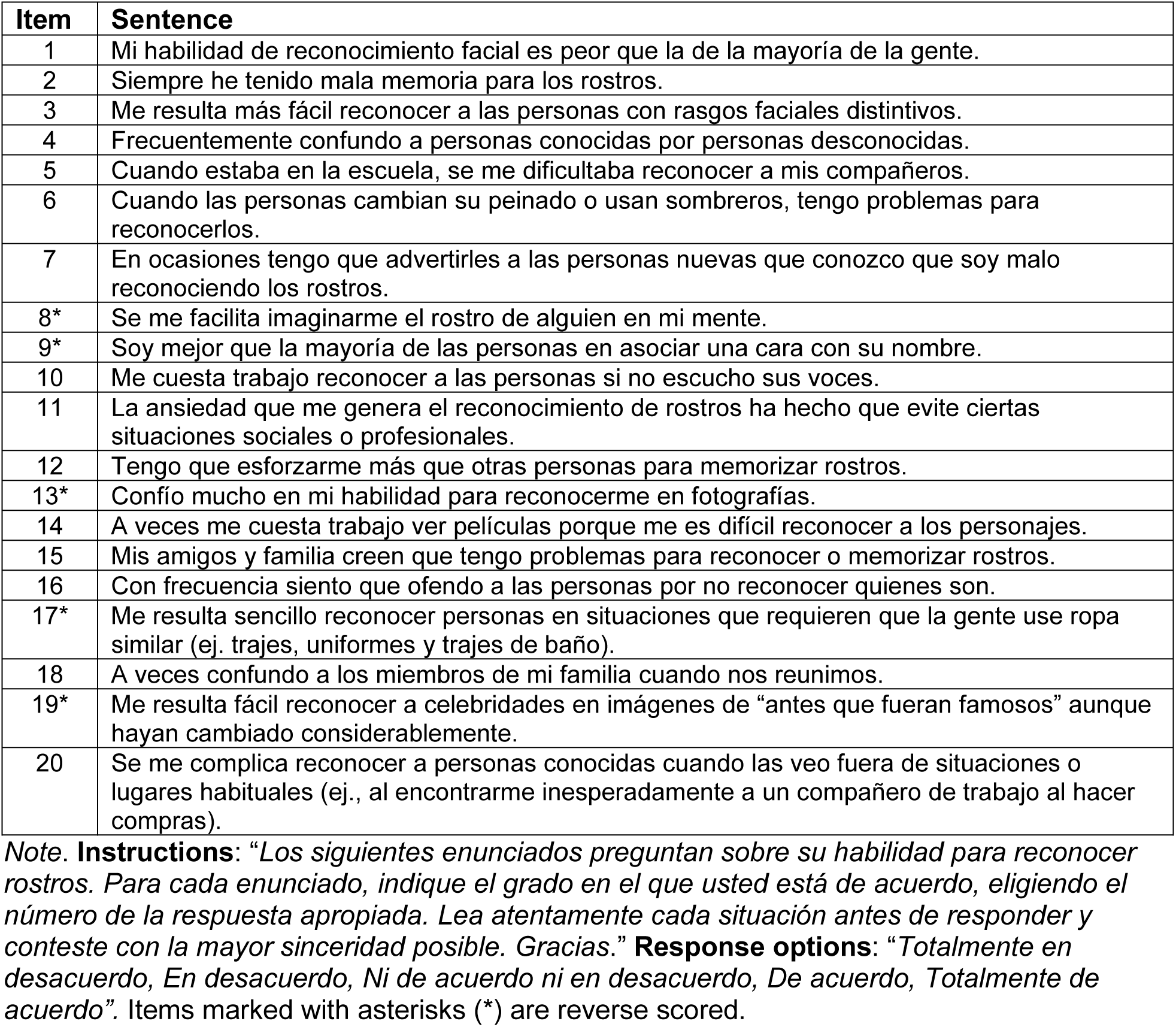
Spanish version of The 20-Item Prosopagnosia Index Items (PI20).

**Tabla S2.** Confirmatory factor analysis of the Spanish version of the PI20 (Study 2).

| Factor | Indicator | Estimate | Std. Error | Z-value | P | 95% Confidence Interval |  |
| --- | --- | --- | --- | --- | --- | --- | --- |
|  |  |  |  |  |  | Lower Range | Upper Range |
| Factor 1 | PI20_01 | 0.686 | 0.018 | 39.205 | < .001 | 0.652 | 0.721 |
|  | PI20_02 | 0.719 | 0.017 | 41.361 | < .001 | 0.685 | 0.753 |
|  | PI20_03 | 0.061 | 0.022 | 2.801 | 0.005 | 0.018 | 0.103 |
|  | PI20_04 | 0.654 | 0.019 | 34.139 | < .001 | 0.617 | 0.692 |
|  | PI20_05 | 0.738 | 0.019 | 39.602 | < .001 | 0.701 | 0.774 |
|  | PI20_06 | 0.689 | 0.018 | 37.809 | < .001 | 0.653 | 0.725 |
|  | PI20_07 | 0.815 | 0.017 | 48.924 | < .001 | 0.783 | 0.848 |
|  | PI20_08 | 0.222 | 0.02 | 11.009 | < .001 | 0.183 | 0.262 |
|  | PI20_09 | 0.298 | 0.021 | 14.481 | < .001 | 0.258 | 0.339 |
|  | PI20_10 | 0.603 | 0.019 | 31.948 | < .001 | 0.566 | 0.64 |
|  | PI20_11 | 0.664 | 0.018 | 36.019 | < .001 | 0.628 | 0.7 |
|  | PI20_12 | 0.756 | 0.017 | 43.966 | < .001 | 0.722 | 0.789 |
|  | PI20_13 | 0.408 | 0.02 | 19.99 | < .001 | 0.368 | 0.448 |
|  | PI20_14 | 0.511 | 0.021 | 24.89 | < .001 | 0.471 | 0.551 |
|  | PI20_15 | 0.686 | 0.019 | 36.813 | < .001 | 0.649 | 0.722 |
|  | PI20_16 | 0.683 | 0.017 | 41.39 | < .001 | 0.651 | 0.715 |
|  | PI20_17 | 0.25 | 0.02 | 12.606 | < .001 | 0.211 | 0.289 |
|  | PI20_18 | 0.606 | 0.022 | 27.161 | < .001 | 0.563 | 0.65 |
|  | PI20_19 | 0.083 | 0.022 | 3.856 | < .001 | 0.041 | 0.126 |
|  | PI20_20 | 0.551 | 0.019 | 28.567 | < .001 | 0.513 | 0.589 |

## Notes

### Competing Interest Statement

The authors have declared no competing interest.

### Author Declarations

Ethical approval was obtained from the Research Ethics Committee of CETYS University (#D-CEI154).

## References

Anzellotti, S., & Caramazza, A. (2014). The neural mechanisms for the recognition of face identity in humans. Frontiers in Psychology, 5. 10.3389/fpsyg.2014.00672

Arifin, W. N. (2025). Sample size calculator. http://wnarifin.github.io

Arizpe, J. M., Saad, E., Douglas, A. O., Germine, L., Wilmer, J. B., & DeGutis, J. M. (2019). Self-reported face recognition is highly valid, but alone is not highly discriminative of prosopagnosia-level performance on objective assessments. Behavior Research Methods, 51(3), 1102–1116. 10.3758/s13428-018-01195-w

Benton, A., Sivan, A. B., Hamsher, K. deS., Varney, N. R., & Spreen, O. (1994). Contributions to Neuropsychological Assessment: A clinical manual (2nd ed.). Oxford University Press.

Bonett, D. G., & Wright, T. A. (2015). Cronbach’s alpha reliability: Interval estimation, hypothesis testing, and sample size planning. Journal of Organizational Behavior, 36(1), 3–15. 10.1002/job.1960

Brewer, R., Gray, K., & Cook, R. (2018). Should developmental prosopagnosia, developmental body agnosia, and developmental object agnosia be considered independent neurodevelopmental conditions? Cognitive Neuropsychology, 35, 59–62. 10.1080/02643294.2018.1433153

Champely, S. (2020). pwr: Basic Functions for Power Analysis (Version 1.3-0) [Computer software]. https://CRAN.R-project.org/package=pwr

Corrow, S. L., Dalrymple, K. A., & Barton, J. J. S. (2016). Prosopagnosia: Current perspectives. Eye and Brain, 8, 165–175. 10.2147/EB.S92838

DeGutis, J., Bahierathan, K., Barahona, K., Lee, E., Evans, T. C., Shin, H. M., Mishra, M., Likitlersuang, J., & Wilmer, J. B. (2023). What is the prevalence of developmental prosopagnosia? An empirical assessment of different diagnostic cutoffs. Cortex, 161, 51–64. 10.1016/j.cortex.2022.12.014

Dennett, H. W., McKone, E., Tavashmi, R., Hall, A., Pidcock, M., Edwards, M., & Duchaine, B. (2012). The Cambridge Car Memory Test: A task matched in format to the Cambridge Face Memory Test, with norms, reliability, sex differences, dissociations from face memory, and expertise effects. Behavior Research Methods, 44(2), 587–605. 10.3758/s13428-011-0160-2

Duchaine, B., & Nakayama, K. (2006). Developmental prosopagnosia: A window to content-specific face processing. Current Opinion in Neurobiology, 16(2), 166–173. 10.1016/j.conb.2006.03.003

Garrido, L., Duchaine, B., & Nakayama, K. (2008). Face detection in normal and prosopagnosic individuals. Journal of Neuropsychology, 2(1), 119–140. 10.1348/174866407X246843

Gignac, G. E., & Szodorai, E. T. (2016). Effect size guidelines for individual differences researchers. Personality and Individual Differences, 102, 74–78. 10.1016/j.paid.2016.06.069

Gray, K. L. H., Bird, G., & Cook, R. (2017). Robust associations between the 20-item prosopagnosia index and the Cambridge Face Memory Test in the general population. Royal Society Open Science, 4(3), 160923. 10.1098/rsos.160923

Hambleton, R. K., & Lee, M. K. (2013). Methods for Translating and Adapting Tests to Increase Cross-Language Validity. In D. H. Saklofske, C. R. Reynolds, & V. Schwean (Eds.), The Oxford Handbook of Child Psychological Assessment (Vol. 1). Oxford University Press. 10.1093/oxfordhb/9780199796304.013.0008

Hu, L., & Bentler, P. M. (1999). Cutoff criteria for fit indexes in covariance structure analysis: Conventional criteria versus new alternatives. Structural Equation Modeling: A Multidisciplinary Journal, 6(1), 1–55. 10.1080/10705519909540118

International Test Commission. (2017). ITC Guidelines for Translating and Adapting Tests (2nd Ed.) (Version 2.4). International Test Commission. www.InTestCom.org

JASP Team. (2025). JASP (Version 0.95.1)[Computer software]. https://jasp-stats.org/

Kim, K. H. (2005). The Relation Among Fit Indexes, Power, and Sample Size in Structural Equation Modeling. Structural Equation Modeling: A Multidisciplinary Journal, 12(3), 368–390. 10.1207/s15328007sem1203_2

Koo, T. K., & Li, M. Y. (2016). A Guideline of Selecting and Reporting Intraclass Correlation Coefficients for Reliability Research. Journal of Chiropractic Medicine, 15(2), 155–163. 10.1016/j.jcm.2016.02.012

Li, C.-H. (2016). Confirmatory factor analysis with ordinal data: Comparing robust maximum likelihood and diagonally weighted least squares. Behavior Research Methods, 48(3), 936–949. 10.3758/s13428-015-0619-7

Nakashima, S. F., Ukezono, M., Sudo, R., Nunoi, M., Kitagami, S., Okubo, M., Toriyama, R., Morimoto, Y., & Takano, Y. (2020). Development of a Japanese version of the 20-item prosopagnosia index (PI20-J) and examination of its reliability and validity. The Japanese Journal of Psychology, 90(6), 603–613. 10.4992/jjpsy.90.18235

Nigrou, T., Hansenne, M., & Devue, C. (2024a). Exploration of the Links Between Psychosocial Well-being and Face Recognition Skills in a French-Speaking Sample. Psychologica Belgica. 10.5334/pb.1294

Nigrou, T., Hansenne, M., & Devue, C. (2024b). Exploration of the Links Between Psychosocial Well-being and Face Recognition Skills in a French-Speaking Sample. Psychologica Belgica, 64(1). 10.5334/pb.1294

Nørkær, E., Guðbjörnsdóttir, E., Roest, S. B., Shah, P., Gerlach, C., & Starrfelt, R. (2023). The Danish Version of the 20-Item Prosopagnosia Index (PI20): Translation, Validation and a Link to Face Perception. Brain Sciences, 13(2), 337. 10.3390/brainsci13020337

Posit team. (2023). RStudio: Integrated Development Environment for R. Posit. http://www.posit.co/.

Sexton, O., Pilley, S., d’Ardenne, J., & Bull, R. (2023). Cognitive Interviewing and what it can be used for. https://www.ncrm.ac.uk/resources/online/all/?id=20816

Shah, P., Gaule, A., Sowden, S., Bird, G., & Cook, R. (2015). The 20-item prosopagnosia index (PI20): A self-report instrument for identifying developmental prosopagnosia. Royal Society Open Science, 2(6), 140343. 10.1098/rsos.140343

Smith, C., & Susilo, T. (2021). Normal colour perception in developmental prosopagnosia. Scientific Reports, 11(1), 13741–13741. 10.1038/s41598-021-92840-6

Stensen, K., & Lydersen, S. (2022). Internal consistency: From alpha to omega? Tidsskrift for Den Norske Legeforening. 10.4045/tidsskr.22.0112

Stumps, A., Saad, E., Rothlein, D., Verfaellie, M., & DeGutis, J. (2020). Characterizing developmental prosopagnosia beyond face perception: Impaired recollection but intact familiarity recognition. Cortex, 130, 64–77. 10.1016/j.cortex.2020.04.016

Sun, W., Wang, Y., Wang, J., & Luo, F. (2021). Psychometric Properties of the Chinese version of the 20-item Prosopagnosia Index (PI20). E3S Web of Conferences, 271, 01036. 10.1051/e3sconf/202127101036

Tagliente, S., Passarelli, M., D’Elia, V., Palmisano, A., Dunn, J. D., Masini, M., Lanciano, T., Curci, A., & Rivolta, D. (2023). Self-reported face recognition abilities moderately predict face-learning skills: Evidence from Italian samples. Heliyon, 9(3), e14125. 10.1016/j.heliyon.2023.e14125

Towler, J., Fisher, K., & Eimer, M. (2017). The cognitive and neural basis of developmental prosopagnosia. Quarterly Journal of Experimental Psychology, 70(2), 316–344. 10.1080/17470218.2016.1165263

Ventura, P., Livingston, L. A., & Shah, P. (2018a). Adults have moderate-to-good insight into their face recognition ability: Further validation of the 20-item Prosopagnosia Index in a Portuguese sample. Quarterly Journal of Experimental Psychology, 71(12), 2677–2679. 10.1177/1747021818765652

Ventura, P., Livingston, L. A., & Shah, P. (2018b). Adults have moderate-to-good insight into their face recognition ability: Further validation of the 20-item Prosopagnosia Index in a Portuguese sample. Quarterly Journal of Experimental Psychology, 71(12), 2677–2679. 10.1177/1747021818765652

World Medical Association Declaration of Helsinki: Ethical Principles for Medical Research Involving Human Subjects. (2013). JAMA, 310(20), 2191. 10.1001/jama.2013.281053

